# The Role of Sepsis Care in Rural Emergency Departments: A Qualitative Study of Emergency Department User Perspectives

**DOI:** 10.1101/2024.05.05.24306891

**Authors:** Nicholas M. Mohr, Kimberly A.S. Merchant, Brian M. Fuller, Brett Faine, Luke Mack, Amanda Bell, Katie DeJong, Edith A. Parker, Keith Mueller, Elizabeth Chrischilles, Christopher R. Carpenter, Michael P. Jones, Steven Q. Simpson, Marcia M. Ward

**Author notes:** **Corresponding Author:** Nicholas M. Mohr, MD, MS, Department of Emergency Medicine, University of Iowa Carver College of Medicine, 200 Hawkins Drive, GH SE203, Iowa City, IA 52242.

## Abstract

**Objective:** Sepsis is a leading cause of hospitalization and death in the United States, and rural patients are at particularly high risk. Telehealth has been proposed as one strategy to narrow rural-urban disparities. The objective of this study was to understand why staff use provider-to-provider telehealth in rural emergency departments (tele-ED) and how tele-ED care changes the care for rural patients with sepsis.

**Methods:** We conducted a qualitative interview study between March 1, 2022 and May 22, 2023 with participants from upper Midwest rural EDs the tele-ED hub physicians in a single tele-ED network that delivers provider-to-provider consultation for sepsis patients. One interviewer conducted individual telephone interviews, then we used standard qualitative methods based on modified grounded theory to identify themes and domains.

**Results:** We interviewed 27 participants, and from the interviews we identified nine themes within three domains. Participants largely felt tele-ED for sepsis was valuable in their practice. We identified that telehealth was consulted to facilitate interhospital transfer, provide surge capacity for small teams, to adhere with policy around provider scope of practice, for inexperienced providers, and for patients with increased severity of illness or complex comorbidities. Barriers to tele-ED use and impact of tele-ED included increased sepsis care standardization, provider reluctance, and sepsis diagnostic uncertainty. Additionally, we identified that real-time education and training were important secondary benefits identified from tele-ED use.

**Conclusions:** Tele-ED care was used by rural providers for sepsis treatment, but many barriers existed that may have limited potential benefits to its use.

## INTRODUCTION

### Background

Sepsis is a leading cause of hospitalization and in-hospital death in the U.S., and patients treated initially in low-volume hospitals have 38% higher mortality than those treated in high-volume centers.^1–3^ Sepsis mortality has fallen over the last 25 years, but care in rural hospitals continues to offer opportunities for improvement.^4–9^ The *Surviving Sepsis Campaign* publishes practice guidelines to recommend care for patients diagnosed with sepsis, but despite those recommendations being incorporated into publicly reported quality metrics, adherence remains imperfect.^10–13^

Provider-to-provider telehealth has been seen as one strategy to provide expert recommendations at the point of care.^14, 15^ Sepsis telehealth applications have been developed for patients in intensive care units, inpatient wards, rural emergency departments (EDs), and post-acute care, and sepsis telehealth in some settings has been associated with improved clinical outcomes.^16–19^

### Importance

The TELEmedicine as a Virtual Intervention for Sepsis in Emergency Departments (TELEVISED) study was developed to understand how real-time video consultation is associated with improved guideline adherence and clinical outcomes in rural patients with sepsis.^20, 21^ An early study showed that telehealth in rural EDs was associated with higher sepsis bundle adherence (adjusted odds ratio [aOR] 17.3, 95% confidence interval [CI] 6.6 to 44.9)^22^, but we subsequently found that one significant barrier to telehealth use was sepsis diagnosis and recognition.^23^ In another multicenter study in telehealth-capable hospitals with contemporary controls (n=1,191), we found telehealth consultation was not associated with improved outcomes, but the subgroup of patients treated in the most remote hospitals by advanced practice providers may have had reduced mortality (aOR 0.11, 95% CI 0.02 to 0.73).^24^ Since these findings were unexpected, we conducted a qualitative study of rural hospital and hub healthcare staff to better understand and interpret the findings from our quantitative studies.

### Goals of This Investigation

The objective of this study was to understand how and why provider-to-provider telehealth was used to care for patients with sepsis in rural EDs and how telehealth recommendations were incorporated into the care and clinical outcomes of rural sepsis patients.

## METHODS

This study was a qualitative analysis of a mixed-methods study using interviews from rural physicians and advanced practice providers (APPs, including physician assistants and nurse practitioners), rural nurses, and telehealth hub physicians from sites in the original TELEVISED cohort. Our mixed methods study used an explanatory sequential design, in which the qualitative interview guide was informed by the results of the quantitative study to add explanation and context to our findings.^25^ For our qualitative design, we used inductive content analysis based on modified grounded theory to identify themes related to each interview topic from interview transcripts.^26^ This study was approved by the local institutional review board, written consent was obtained, and our findings are reported in accordance with the Consolidated Criteria for Reporting Qualitative Research (COREQ).^27^ Methodologic details are included **Supplemental Appendix A**.

### Intervention, Participants, and Interviews

We conducted our interviews among sites that participate in a single hub-and-spoke provider-to-provider emergency department-based telehealth (tele-ED) network based in Sioux Falls, South Dakota. That network provides on-demand high-definition video consultation to 214 rural EDs across 13 states, 24 hours daily with board-certified emergency physicians and experienced nurses at the hub. Potential participants were identified by local medical staff, and interviews were conducted using a semi-structured interview guide (**Supplemental Appendix B**) between March 1, 2022 and May 22, 2023.

### Qualitative Analysis

After interviews, two coders independently reviewed all transcripts and developed a qualitative codebook based upon the themes that emerged in the interviews. Themes were identified in domains based on discussion within the study team, and disagreements were resolved by discussion with a third qualitative analyst. The codebook and coding were performed using Microsoft Word and Microsoft Excel (Microsoft Corporation, Redmond, Washington), and codes and themes were not shared with participants. The major domains and themes were presented with illustrative quotations, and the study team discussed data interpretation iteratively to ensure consistency.

## RESULTS

We enrolled a total of 27 participants from 13 hospitals: five hub physicians, eight rural ED providers (four physicians and four APPs, from eight unique hospitals), and 14 rural nurses (from 11 unique hospitals). Rural participants varied in use of tele-ED based on scope of practice, experience, and comfort with both telehealth and sepsis care (**Table 1**).

**Table 1.** Characteristics of study participants.

| **Characteristic** | **n (%)**<br>**(n=27)** |
| --- | --- |
| Professional Role |  |
| Hub Physician | 5 (19) |
| Rural ED Provider |  |
| Physician | 4 (15) |
| Advanced Practice Provider | 4 (15) |
| Nurse | 14 (52) |
| Years of Telehealth Experience (Rural Staff Only) |  |
| 0-5 | 4 (18) |
| 6-10 | 7 (32) |
| 11-15 | 5 (23) |
| 16 or greater | 1 (5) |
| Unknown | 5 (23) |
| Years of Telehealth Hub Experience |  |
| Less than 5 | 2 (40) |
| More than 10 | 3 (60) |
| Proportion of Sepsis Telehealth Use (Rural Staff Only) |  |
| 0-25% | 7 (32) |
| 26-50% | 6 (27) |
| 51-75% | 4 (18) |
| 76-100% | 4 (18) |
| Unknown | 1 (5) |
| Hospital Designation (Rural Staff Only) |  |
| Critical Access | 19 (86) |
| Prospective Payment System | 3 (14) |

Interview responses were organized into three main domains: (1) facilitators and benefits to using tele-ED; (2) barriers and factors that mitigate potential benefits of tele-ED use; and (3) other considerations associated with tele-ED use for sepsis. Within the first two domains, we identified themes focused on (1) hospital/facility factors, (2) staff factors, and (3) patient factors. **Figure 1** presents the conceptual model of our qualitative findings, and **Table 2** describes themes with exemplary quotes.

**Figure 1.**
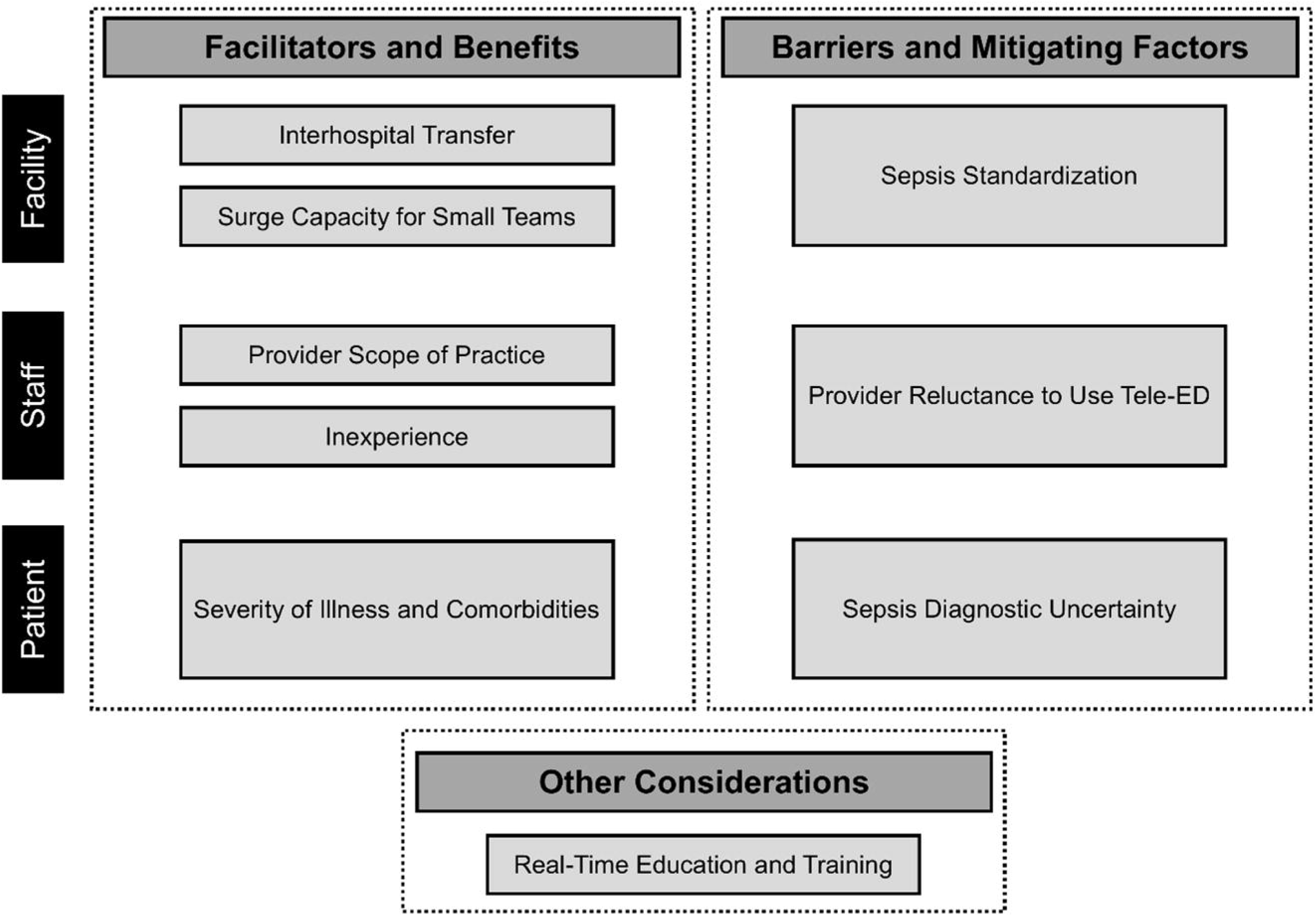
Themes and domains of qualitative findings. We identified 9 themes from our qualitative data that grouped into 3 main domains: facilitators and benefits of tele-emergency (tele-ED) use, barriers and factors mitigating clinical benefits of tele-ED use, and other considerations. Within the first 2 categories, we considered themes according to three categories: facility-based, staff-based, and patient-based themes.

**Table 2.**
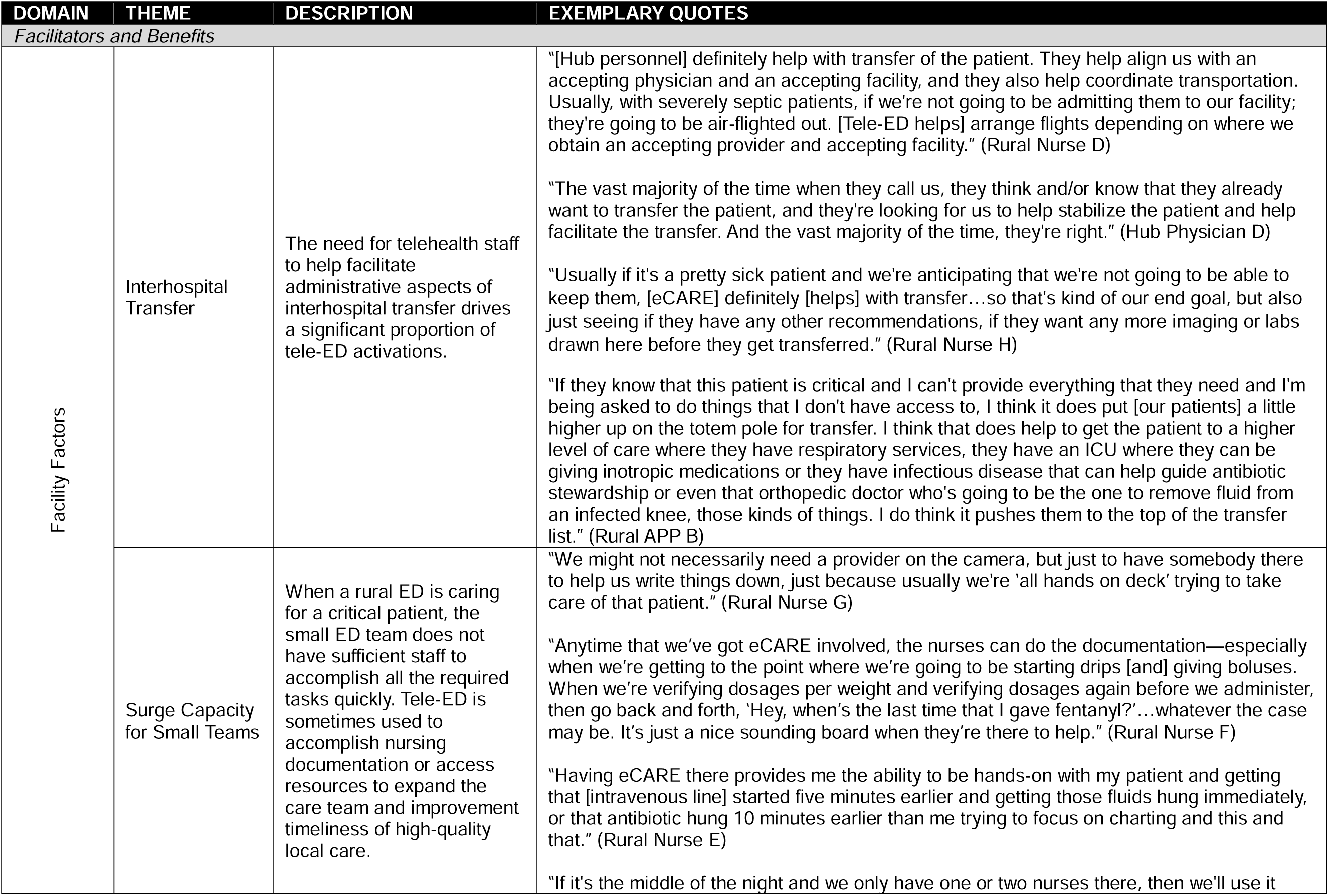
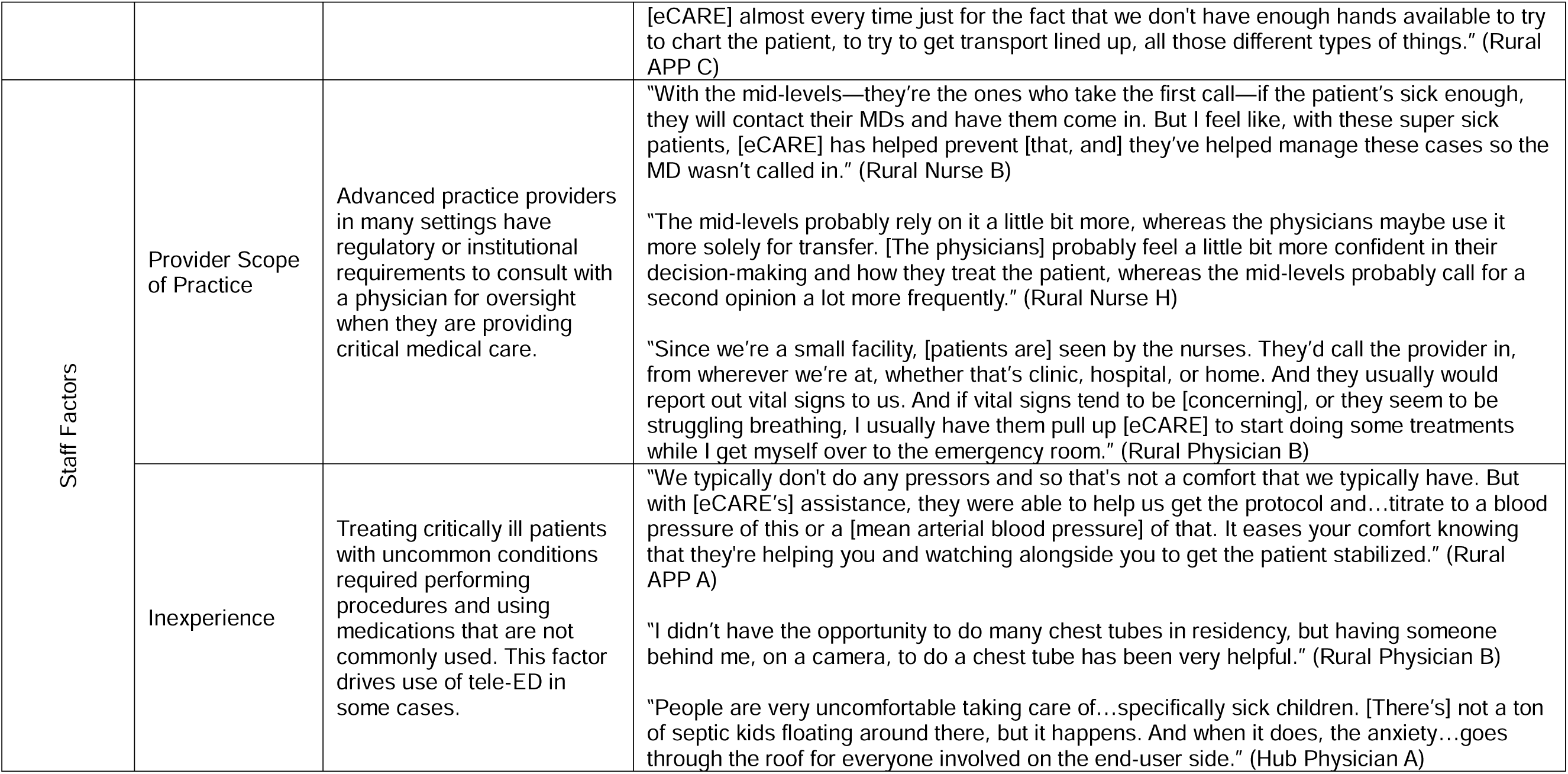
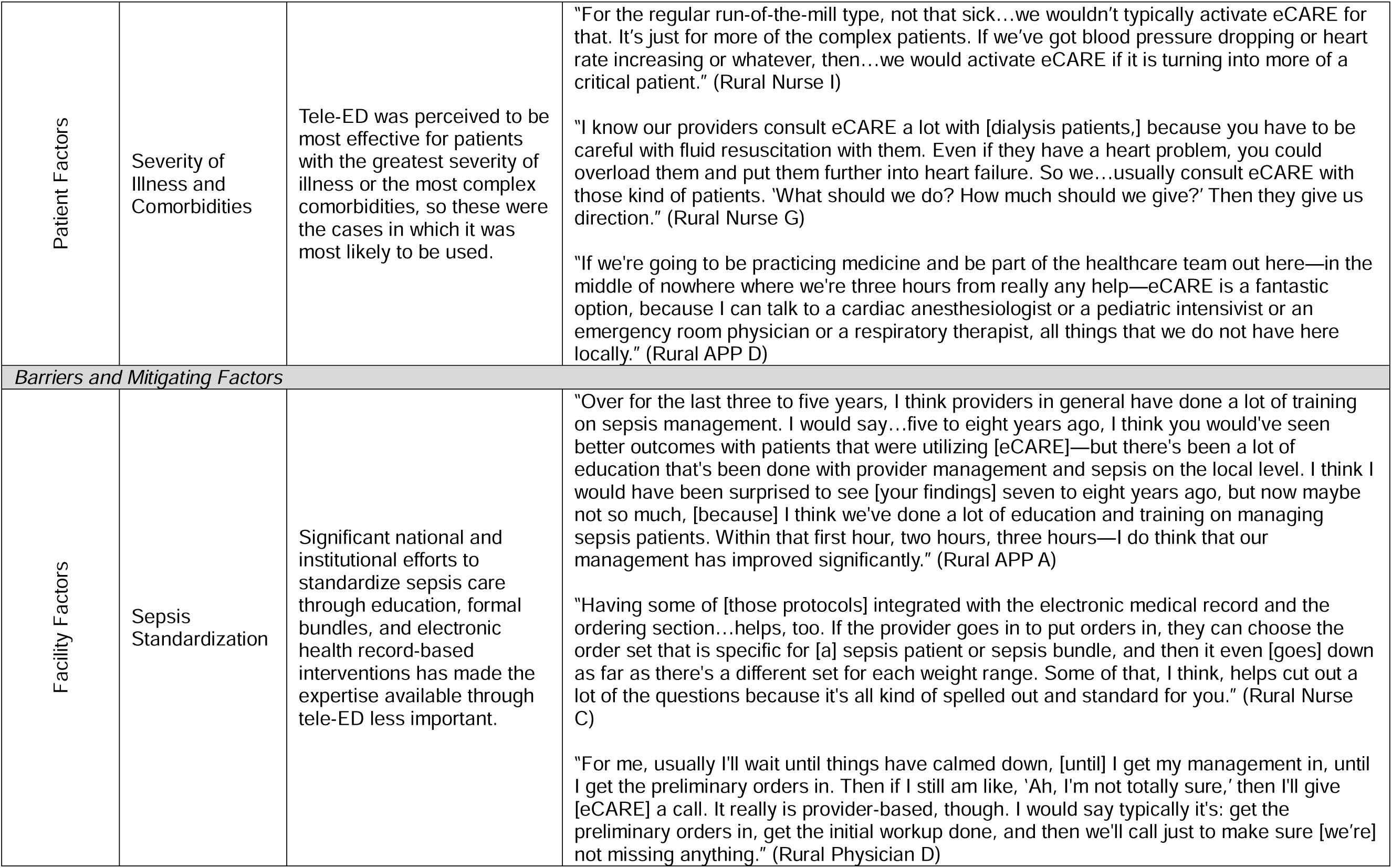
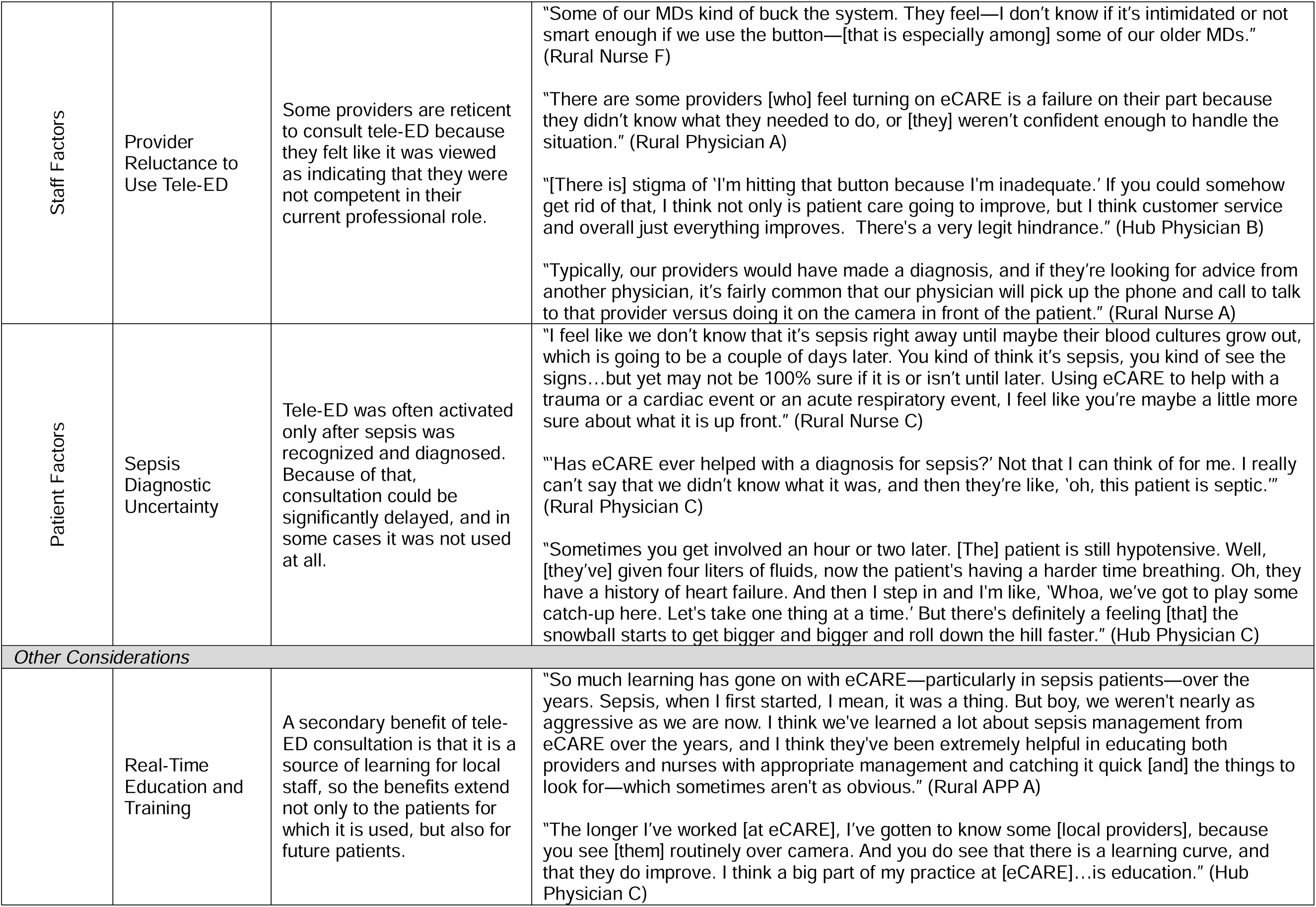
Qualitative themes and exemplary quotes related to use of provider-to-provider emergency department telehealth for rural sepsis patients. *APP, advanced practice provider*

| DOMAIN | THEME | DESCRIPTION | EXEMPLARY QUOTES |
| --- | --- | --- | --- |
| *Facilitators and Benefits* |  |  |  |
| Facility Factors | Interhospital Transfer | The need for telehealth staff to help facilitate administrative aspects of interhospital transfer drives a significant proportion of tele-ED activations. | “[Hub personnel] definitely help with transfer of the patient. They help align us with an accepting physician and an accepting facility, and they also help coordinate transportation. Usually, with severely septic patients, if we’re not going to be admitting them to our facility; they’re going to be air-flighted out. [Tele-ED helps] arrange flights depending on where we obtain an accepting provider and accepting facility.” (Rural Nurse D)<br>“The vast majority of the time when they call us, they think and/or know that they already want to transfer the patient, and they’re looking for us to help stabilize the patient and help facilitate the transfer. And the vast majority of the time, they’re right.” (Hub Physician D)<br>“Usually if it’s a pretty sick patient and we’re anticipating that we’re not going to be able to keep them, [eCARE] definitely [helps] with transfer...so that’s kind of our end goal, but also just seeing if they have any other recommendations, if they want any more imaging or labs drawn here before they get transferred.” (Rural Nurse H)<br>“If they know that this patient is critical and I can’t provide everything that they need and I’m being asked to do things that I don’t have access to, I think it does put [our patients] a little higher up on the totem pole for transfer. I think that does help to get the patient to a higher level of care where they have respiratory services, they have an ICU where they can be giving inotropic medications or they have infectious disease that can help guide antibiotic stewardship or even that orthopedic doctor who’s going to be the one to remove fluid from an infected knee, those kinds of things. I do think it pushes them to the top of the transfer list.” (Rural APP B) |
|  | Surge Capacity for Small Teams | When a rural ED is caring for a critical patient, the small ED team does not have sufficient staff to accomplish all the required tasks quickly. Tele-ED is sometimes used to accomplish nursing documentation or access resources to expand the care team and improvement timeliness of high-quality local care. | “We might not necessarily need a provider on the camera, but just to have somebody there to help us write things down, just because usually we’re ‘all hands on deck’ trying to take care of that patient.” (Rural Nurse G)<br>“Anytime that we’ve got eCARE involved, the nurses can do the documentation—especially when we’re getting to the point where we’re going to be starting drips [and] giving boluses. When we’re verifying dosages per weight and verifying dosages again before we administer, then go back and forth, ‘Hey, when’s the last time that I gave fentanyl?’...whatever the case may be. It’s just a nice sounding board when they’re there to help.” (Rural Nurse F)<br>“Having eCARE there provides me the ability to be hands-on with my patient and getting that [intravenous line] started five minutes earlier and getting those fluids hung immediately, or that antibiotic hung 10 minutes earlier than me trying to focus on charting and this and that.” (Rural Nurse E)<br>“If it’s the middle of the night and we only have one or two nurses there, then we’ll use it |
|  |  |  | [eCARE] almost every time just for the fact that we don't have enough hands available to try to chart the patient, to try to get transport lined up, all those different types of things." (Rural APP C) |
| Staff Factors | Provider Scope of Practice | Advanced practice providers in many settings have regulatory or institutional requirements to consult with a physician for oversight when they are providing critical medical care. | "With the mid-levels—they're the ones who take the first call—if the patient's sick enough, they will contact their MDs and have them come in. But I feel like, with these super sick patients, [eCARE] has helped prevent [that, and] they've helped manage these cases so the MD wasn't called in." (Rural Nurse B)<br>"The mid-levels probably rely on it a little bit more, whereas the physicians maybe use it more solely for transfer. [The physicians] probably feel a little bit more confident in their decision-making and how they treat the patient, whereas the mid-levels probably call for a second opinion a lot more frequently." (Rural Nurse H)<br>"Since we're a small facility, [patients are] seen by the nurses. They'd call the provider in, from wherever we're at, whether that's clinic, hospital, or home. And they usually would report out vital signs to us. And if vital signs tend to be [concerning], or they seem to be struggling breathing, I usually have them pull up [eCARE] to start doing some treatments while I get myself over to the emergency room." (Rural Physician B) |
|  | Inexperience | Treating critically ill patients with uncommon conditions required performing procedures and using medications that are not commonly used. This factor drives use of tele-ED in some cases. | "We typically don't do any pressors and so that's not a comfort that we typically have. But with [eCARE's] assistance, they were able to help us get the protocol and...titrate to a blood pressure of this or a [mean arterial blood pressure] of that. It eases your comfort knowing that they're helping you and watching alongside you to get the patient stabilized." (Rural APP A)<br>"I didn't have the opportunity to do many chest tubes in residency, but having someone behind me, on a camera, to do a chest tube has been very helpful." (Rural Physician B)<br>"People are very uncomfortable taking care of...specifically sick children. [There's] not a ton of septic kids floating around there, but it happens. And when it does, the anxiety...goes through the roof for everyone involved on the end-user side." (Hub Physician A) |
| Patient Factors | Severity of Illness and Comorbidities | Tele-ED was perceived to be most effective for patients with the greatest severity of illness or the most complex comorbidities, so these were the cases in which it was most likely to be used. | “For the regular run-of-the-mill type, not that sick...we wouldn’t typically activate eCARE for that. It’s just for more of the complex patients. If we’ve got blood pressure dropping or heart rate increasing or whatever, then...we would activate eCARE if it is turning into more of a critical patient.” (Rural Nurse I)<br>“I know our providers consult eCARE a lot with [dialysis patients,] because you have to be careful with fluid resuscitation with them. Even if they have a heart problem, you could overload them and put them further into heart failure. So we...usually consult eCARE with those kind of patients. ‘What should we do? How much should we give?’ Then they give us direction.” (Rural Nurse G)<br>“If we’re going to be practicing medicine and be part of the healthcare team out here—in the middle of nowhere where we’re three hours from really any help—eCARE is a fantastic option, because I can talk to a cardiac anesthesiologist or a pediatric intensivist or an emergency room physician or a respiratory therapist, all things that we do not have here locally.” (Rural APP D) |
| *Barriers and Mitigating Factors* |  |  |  |
| Facility Factors | Sepsis Standardization | Significant national and institutional efforts to standardize sepsis care through education, formal bundles, and electronic health record-based interventions has made the expertise available through tele-ED less important. | “Over for the last three to five years, I think providers in general have done a lot of training on sepsis management. I would say...five to eight years ago, I think you would’ve seen better outcomes with patients that were utilizing [eCARE]—but there’s been a lot of education that’s been done with provider management and sepsis on the local level. I think I would have been surprised to see [your findings] seven to eight years ago, but now maybe not so much, [because] I think we’ve done a lot of education and training on managing sepsis patients. Within that first hour, two hours, three hours—I do think that our management has improved significantly.” (Rural APP A)<br>“Having some of [those protocols] integrated with the electronic medical record and the ordering section...helps, too. If the provider goes in to put orders in, they can choose the order set that is specific for [a] sepsis patient or sepsis bundle, and then it even [goes] down as far as there’s a different set for each weight range. Some of that, I think, helps cut out a lot of the questions because it’s all kind of spelled out and standard for you.” (Rural Nurse C)<br>“For me, usually I’ll wait until things have calmed down, [until] I get my management in, until I get the preliminary orders in. Then if I still am like, ‘Ah, I’m not totally sure,’ then I’ll give [eCARE] a call. It really is provider-based, though. I would say typically it’s: get the preliminary orders in, get the initial workup done, and then we’ll call just to make sure [we’re] not missing anything.” (Rural Physician D) |
| Staff Factors | Provider Reluctance to Use Tele-ED | Some providers are reticent to consult tele-ED because they felt like it was viewed as indicating that they were not competent in their current professional role. | “Some of our MDs kind of buck the system. They feel—I don’t know if it’s intimidated or not smart enough if we use the button—[that is especially among] some of our older MDs.” (Rural Nurse F)<br>“There are some providers [who] feel turning on eCARE is a failure on their part because they didn’t know what they needed to do, or [they] weren’t confident enough to handle the situation.” (Rural Physician A)<br>“[There is] stigma of ‘I’m hitting that button because I’m inadequate.’ If you could somehow get rid of that, I think not only is patient care going to improve, but I think customer service and overall just everything improves. There’s a very legit hindrance.” (Hub Physician B)<br>“Typically, our providers would have made a diagnosis, and if they’re looking for advice from another physician, it’s fairly common that our physician will pick up the phone and call to talk to that provider versus doing it on the camera in front of the patient.” (Rural Nurse A) |
| Patient Factors | Sepsis Diagnostic Uncertainty | Tele-ED was often activated only after sepsis was recognized and diagnosed. Because of that, consultation could be significantly delayed, and in some cases it was not used at all. | “I feel like we don’t know that it’s sepsis right away until maybe their blood cultures grow out, which is going to be a couple of days later. You kind of think it’s sepsis, you kind of see the signs...but yet may not be 100% sure if it is or isn’t until later. Using eCARE to help with a trauma or a cardiac event or an acute respiratory event, I feel like you’re maybe a little more sure about what it is up front.” (Rural Nurse C)<br>“‘Has eCARE ever helped with a diagnosis for sepsis?’ Not that I can think of for me. I really can’t say that we didn’t know what it was, and then they’re like, ‘oh, this patient is septic.’” (Rural Physician C)<br>“Sometimes you get involved an hour or two later. [The] patient is still hypotensive. Well, [they’ve] given four liters of fluids, now the patient’s having a harder time breathing. Oh, they have a history of heart failure. And then I step in and I’m like, ‘Whoa, we’ve got to play some catch-up here. Let’s take one thing at a time.’ But there’s definitely a feeling [that] the snowball starts to get bigger and bigger and roll down the hill faster.” (Hub Physician C) |
| *Other Considerations* |  |  |  |
|  | Real-Time Education and Training | A secondary benefit of tele-ED consultation is that it is a source of learning for local staff, so the benefits extend not only to the patients for which it is used, but also for future patients. | “So much learning has gone on with eCARE—particularly in sepsis patients—over the years. Sepsis, when I first started, I mean, it was a thing. But boy, we weren’t nearly as aggressive as we are now. I think we’ve learned a lot about sepsis management from eCARE over the years, and I think they’ve been extremely helpful in educating both providers and nurses with appropriate management and catching it quick [and] the things to look for—which sometimes aren’t as obvious.” (Rural APP A)<br>“The longer I’ve worked [at eCARE], I’ve gotten to know some [local providers], because you see [them] routinely over camera. And you do see that there is a learning curve, and that they do improve. I think a big part of my practice at [eCARE]...is education.” (Hub Physician C) |

### Facilitators and Benefits to Using Tele-ED in Rural ED Sepsis

Factors driving tele-ED use or tele-ED benefits in rural sepsis care include facility factors (interhospital transfer; surge capacity for small teams), staffing and provider factors (provider scope of practice; inexperience), and patient factors (severity of illness and comorbidities).

#### Facility Factor: Interhospital Transfer

Interfacility transfer plays a critical role in whether rural providers activate tele-ED, but reasons for transfer can vary according to local context. Many small rural hospitals have limited inpatient capacity, procedural capabilities, or staffing that preclude local admission, and most hospitals in our study did not have intensive care units. Many participants shared that the need for interhospital transfer was the primary reasons that tele-ED was consulted. Tele-ED providers often completed the administrative tasks for interhospital transfer to free up local ED staff for other work.

In most cases, hub physicians did not determine the need for transfer—this was usually done before tele-ED activation. That factor also contributed to timing of tele-ED use because local staff often waited for diagnostic test results and the assessment of the initial response to therapy before deciding whether transfer was necessary; thus, much of the early medical care was provided prior to tele-ED activation, which contributed to consultation delays.

As part of helping with transfer, however, medical guidance was often provided. Participants varied on their perception of the extent to which treatment recommendations were made—some interactions led to detailed conversations about patient management and others focused mostly on the administrative aspects of transfer themselves, and the character of this conversation was determined by local staff preference. In many cases, however, both hub staff and rural staff highlighted that suggestions, recommendations, and tips were informally provided while they were still in rural EDs.

#### Facility Factor: Surge Capacity for Small Teams

Many participating rural EDs were staffed with small teams: sometimes only a single provider and two nurses who may also be caring for hospital inpatients. While that was often sufficient for routine care, teams were shorthanded when critically ill patients were being treated—and few EDs had a mechanism to marshal additional staff for periods of workload surge. For nurses specifically, tele-ED was often used for documentation or to consult medical references (e.g., drug compatibility, etc.), which was one way to expand the effectiveness of a small clinical team by offloading administrative tasks to hub nurses.

Even during these tele-ED activations, hub staff provided guidance and checked for errors, which improved timeliness of care for sepsis patients. They were able to enter electronic orders and track completion of bundle elements for local staff who continued bedside treatment.

#### Staff Factors: Provider Scope of Practice

Many participants felt that tele-ED consultation was used very differently by different provider groups—partially for formal supervision or because of state or institutional policies. Some observed that APPs were more likely to use tele-ED, and participants thought that was because they valued the experience and training of the tele-ED provider, were more accustomed to talking through care pathways for critically ill patients, or were required by institutional or state policy. APPs acknowledged their comfort with the technology and the interactions with the hub. Supervision requirements sometimes applied to specific types of patients (e.g., critically ill patients, transfer patients), which drove tele-ED use.

Tele-ED was also sometimes activated because no physician or APP was present in the ED. In those cases, a tele-ED physician functioned as the primary individual guiding care without a local provider—usually until a local provider arrived from clinic, elsewhere in the hospital, or from home.

#### Staff Factors: Inexperience with Medications or Procedures

Several participants highlighted that staff in the most remote facilities were more likely to activate tele-ED sooner because they see sepsis cases less frequently—often to provide guidance for rarely used medications or for uncommon procedures. This use was especially frequent for sepsis patients who required endotracheal intubation, in which procedural guidance is a well-established tele-ED practice.

Medication guidance was reported to focus principally on drug selection and dosing. This use was most common for antibiotics, vasopressors, or practical aspects like drug compatibility and infusion rates. Some participants also discussed the frequency of using tele-ED for providing pediatric sepsis care, which was a particularly rare event in all our participants’ hospitals. In these scenarios, tele-ED providers were able to provide a broader experience related to drug selection and dosing, risk stratification, and consulting reference materials as required.

#### Patient Factors: Severity of Illness and Comorbidities

Another theme that persisted about both use and timing of tele-ED activation was the role of illness severity and comorbidities in deciding when to use tele-ED. Those with more severe disease may have more acute medical needs and often required specific or unique medications and procedures less familiar to treating staff. For instance, the most severely ill patients with multiple organ failure more often required complex management that local providers consulted with tele-ED staff to co-manage.

The other situation that encouraged tele-ED use were patients with multiple competing comorbidities—especially congestive heart failure or end-stage renal disease. Several participants also noted that patients with the most severe illness or who had multiple complex conditions were the patients in which they thought that tele-ED recommendations could have the most impact.

### Barriers and Factors that Mitigate Benefits in Rural ED Sepsis

We identified three themes focused on barriers preventing tele-ED use and factors that mitigate potential benefits of tele-ED in rural sepsis patients: standardization of protocols, provider reluctance, and diagnostic uncertainty.

#### Facility Factor: Sepsis Standardization Reduces Variation in Non-Tele-ED Care

Several rural staff members highlighted the important role that sepsis care standardization has played in attenuating beneficial effects of tele-ED. In many cases, this standardization has been reflected in extensive educational initiatives, standard screening protocols, treatment order sets, and performance feedback. Many of our participants felt overall sepsis care had improved over the past 10 years on account of these activities, even without tele-ED use. These initiatives, in their opinion, translated to increased comfort and effectiveness for local staff to manage sepsis cases effectively.

That increased comfort also translated into later tele-ED consultations. Many of the local ED staff felt that following protocols usurped tele-ED consultation, which could be an explanation for the limited tele-ED impact in our quantitative results. Several staff suggested that in a system without the substantial focus on sepsis quality of care, the benefit from tele-ED consultation may have been greater.

#### Staff Factor: Provider Reluctance to Use Tele-ED

One of the consistent themes identified as a barrier to tele-ED use was the reluctance to use a platform in front of staff or patients, because it was viewed as a threat to a rural provider’s professional credibility. This factor was described by rural providers, nurses, and hub physicians, and each group had examples of individual cases where they had seen this as a barrier. Several nurses related stories when they wanted to activate tele-ED and a physician asked them not to do so. Participants explained this behavior as providers feeling confident about their care, not wanting another provider to threaten their professional autonomy, or being concerned about perceptions of inadequacy, but this was a prominent theme.

A factor that many rural providers cited was the importance of respectful and collegial interactions in encouraging future tele-ED use. Few participants had examples of conflict between tele-ED and hub staff, but this seemed to be a concern that several suggested could affect rural staff willingness to use the tele-ED service. Several rural nurses had examples of cases in which rural providers chose to leave the patient room and initiate a telephone consultation (instead of using the video platform), presumably to maintain credibility with local staff, patients, and family members.

#### Patient Factor: Sepsis Diagnostic Uncertainty

This treatment-oriented tele-ED intervention sometimes prevented tele-ED from being consulted, because sepsis often presents with vague symptoms and the diagnosis was sometimes unclear early in the clinical course. Rural participants did not routinely view tele-ED consultation as helping identify patients with sepsis. In fact, they viewed that diagnosis was the rural provider’s role prior to tele-ED activation. Consequently, most of our rural participants did not feel that the tele-ED service contributed significantly to sepsis recognition or diagnosis.

This area was one where rural and hub clinicians held different perspectives. Some hub physicians felt that tele-ED consultation delays were common because of delayed recognition, and that these delays adversely affected the ability for tele-ED to positively influence sepsis treatment.

### Other Considerations Related to Tele-ED in Rural ED Sepsis

We identified one additional consideration that was not cited as a reason for or against tele-ED use, but it was identified by multiple providers as a potential secondary benefit: real-time sepsis education and training.

#### Real-Time Sepsis Education and Training

Some providers noted that learning from having consulted tele-ED in the past contributed to improved future performance. Because of that observation, cases in which tele-ED was not consulted received care more similar to that recommended by tele-ED (from real-time experiential learning). This experiential learning was particularly impactful, because recommendations were provided in a rural staff member’s care context—making it easily transferable to future patients. That method of learning was distinct from didactic lectures, online training modules, continuing education programs, or other alternative methods of ongoing learning: walking through a case in real-time with an expert uniquely contributed to understanding because it allowed local staff to translate knowledge into practical action. When tele-ED was not used, some rural staff indicated that was because local providers were confident with the care they were providing—and that confidence sometimes came from prior tele-ED use.

## LIMITATIONS

This study has several limitations. First, the COVID-19 public health emergency significantly affected operations in participating hospitals. Our quantitative data was from prior to COVID-19, but our interviews were conducted after the pandemic began. We prompted participants to respond based on their experience separate from the public health emergency, but the COVID-19 experience could still have affected their perspectives. Second, our rural participants work in different sizes of hospitals in different types of communities, but they used a single tele-ED service, so the findings may not be fully generalizable to telehealth providers with different structures, functions, or procedures. Finally, the health system from which our participants were recruited has put significant effort into sepsis training and quality improvement over the last 10 years. Some of that effort may have influenced the perceived use and utility of the tele-ED intervention, which may not reflect the experience in other health systems.

## DISCUSSION

In this qualitative follow-up to the original TELEVISED quantitative analysis^24^, we found that telehealth was used most frequently—and was perceived to have the greatest value— in very discrete clinical scenarios. Patients requiring interhospital transfer, those treated by less experienced providers or who had regulatory requirement-mandated supervision were those for whom tele-ED was used most often. Additionally, we identified that standardization of sepsis care, ongoing professional education, and diagnostic uncertainty may have mitigated potential tele-ED benefits in our primary analysis. These findings are valuable, because they highlight that the utility of an acute care telehealth intervention is less a function of the utility of the technology or the telehealth team and more a function of the infrastructure and context surrounding provider-to-provider tele-ED use.

Prior studies have examined the perceived utility of provider-to-provider telehealth, and they have largely found telehealth to add perceived value to care.^28^ In a qualitative study of physicians who experienced an ED-based pediatric tele-resuscitation program, physicians found value in the ability of a telehealth provider to help integrate findings, communicate expectations, and address a local lack of trained staff—based partially on the reputation of the remote hospital providing tele-consultation.^29^ Like our study, the authors identified the importance of collegial interactions, but consultations in their network were more frequent for medical consultation, rather than to facilitate administrative or technical aspects of care. A similar study in the Veterans Health Administration identified apprehension about the use of a new process as a potential barrier to use of an emergency tele-psychiatry program, but the perceived value of consultation was the most important factor driving telehealth use.^30^ Real-time learning was previously identified as an ancillary benefit of telehealth consultation in EDs, and we previously demonstrated that care in tele-ED hospitals changes over time in response to telehealth provider recommendations—suggesting that real-time learning may be one of the mechanisms of telehealth benefits.^31–33^ Our findings reinforce that this specific benefit of telehealth may leverage recommendations so that they impact the treatment not only for an individual patient, but also as a vehicle for disseminating changes in care. Finally, a systematic review demonstrated that telehealth may improve outcomes for sepsis patients, which could be a message that drives tele-ED use.^16^ Our findings corroborate that benefits of telehealth consultation may be attenuated in a health system with significant institutional focus on sepsis quality of care.

The findings from this study reinforce something that sepsis researchers already know: a standardized approach to sepsis care is associated with better sepsis outcomes. Participation in a sepsis quality improvement program improves sepsis outcomes.^34, 35^ Care bundles, provider feedback, education, and nurse-initiated order sets can all improve sepsis standardization.^11, 36–38^ In that context, telehealth may be an additional strategy that can effectively standardize treatment, but in a health system with other performance improvement activities ongoing, the incremental impact may be limited.

The issue of context, though, may be relevant. With health systems experiencing decreased experienced long-term staff after the COVID-19 pandemic, regional approaches to sepsis care that can endure despite staffing turnover are valuable.^39, 40^ This observation may also highlight the importance of real-time learning from telehealth providers.^31^ Disseminating practice change is difficult, and it may be even more challenging in rural hospitals across large geographic areas.^41–43^ Changing practice requires applying new knowledge to a local care context, which telehealth-enabled collaborative medical care may be particularly adept at facilitating.^44^ Having reliable systems to disseminate new guidelines, ensure standardization of care for high-risk conditions, and maintain quality during periods of staffing turbulence could be additional roles acute care provider-to-provider telehealth might play.

In conclusion, provider-to-provider telehealth use in rural EDs for patients with sepsis was viewed by local staff as valuable, but many consultations were initiated to facilitate administrative and technical aspects of care. Further, the focus on sepsis treatment after diagnosis and ongoing professional training in a health system with significant sepsis focus may have attenuated the benefits of sepsis treatment we expected to see. Future work will focus on the context in which telehealth consultation may be most valuable and the structure and process of telehealth-augmented care that can maximize the impact on patient outcomes.

## Data Availability

All data produced in the present study are available upon reasonable request to the authors.

## ACKNOWLEDGEMENTS

The authors acknowledge Tera Shea, BS for her editorial assistance in the preparation of this manuscript.

## Institution where the Work was Performed

University of Iowa Carver College of Medicine

## Prior Presentations

This work was presented at the American Telemedicine Association NEXUS 2024 on May 6, 2024 (Phoenix, Arizona) and at the Society for Academic Emergency Medicine Annual Meeting on May 15, 2024 (Phoenix, Arizona).

## Data Sharing Statement

Because interview transcripts are all identifiable, data are not available for sharing.

## Author Contributions

NMM conceived the study, supervised conduct of the study, participated in data analysis and interpretation, acquired funding, drafted the manuscript, and takes overall responsibility for the integrity of the data. KASM conducted interviews, conducted data analysis, participated in data interpretation, and drafted the manuscript. BMF, BF, EAP, KM, EC, CRC, MPJ, and SQS conceived the study, participated in data interpretation, provided methodologic expertise, and critically revised the manuscript for important intellectual content. LM, AB, and KD participated in conduct of the interviews, participated in data interpretation, and critically revised the manuscript for the manuscript for important intellectual content. MMW conceived the study, supervised conduct of the study, provided methodologic expertise, participated in data interpretation, acquired funding, and critically revised the manuscript for important intellectual content.

