## Appendix A for "The Role of Sepsis Care in Rural Emergency Departments: A Qualitative Study of Emergency Department User Perspectives"

**Supplemental Appendix A. Detailed Methods.**

***Intervention***

The emergency department-based telehealth program provides on-demand high-definition video consultation to 214 rural EDs across 13 states, 24 hours daily with board-certified emergency physicians and experienced nurses at the hub. Each of the hub physicians also practices in a tertiary center ED on days when they are not providing tele-ED services. The tele-ED connection can be activated by physicians, APPs, or nurses in participating rural hospitals by pushing a button on the wall, enabling the hub staff to provide medical recommendations, document physician or nursing records, review results in the electronic health record (EHR), place computerized orders, and arrange interhospital transfer if required. All EDs participating in this study were part of the same health system and used the same EHR, and each of these EDs had local providers on-site or on-call to provide in-person care. Hospitals participating in the tele-ED network have standard nurse and provider sepsis education, nurse-directed sepsis screening with EHR-based alerts, and tele-ED consultation was recommended for sepsis patients starting in 2017.[45]

***Participants***

We planned to recruit hub physicians, nurses, and providers (physicians and APPs) across sites that participated in the original TELEVISED comparative effectiveness study (23 hospitals).[24] Prospective participants were identified by local medical directors and nurse managers, then they were invited to participate through a letter from the study team. Participants were eligible only if they had used tele-ED for the care of a patient with sepsis. Participants were diverse according to their training (physicians in a variety of specialties, APPs, and nurses), telehealth experience (newer and veteran users), and hospital designation (critical access hospitals and prospective payment system hospitals). Interviews lasted approximately 45 minutes, and participants were compensated. All participants completed the interview, and no follow-ups or repeat interviews were performed.

***Interviews***

The guide was developed by two experts in telehealth use for sepsis. We elicited structured feedback from health care personnel with experience working in the telehealth network and adjusted the guide accordingly. Separate prompts were used for each category of participants, but they were harmonized to allow comparative analysis between rural physicians/APPs, nurses, and hub staff. Some questions disclosed results from the parent TELEVISED quantitative study, and participants were asked to try to explain why those findings might have been observed. We conducted a pilot test of the interview guide and iteratively revised it based on feedback and experience in use. All interviews were conducted between March 1, 2022 and May 22, 2023 by the same interviewer with a background in journalism, explicit training in the protocol, and extensive experience conducting qualitative interviews focused on rural telehealth (KASM). None of the participants knew the interviewer prior to the study. Interviews were conducted by telephone individually (i.e., no one other than a single participant, the interviewer, and a study investigator [only on hub participant interviews] were on the call). Participants provided verbal informed consent, and the study team did not disclose to employers or colleagues whether prospective participants agreed to enroll. Interviews were recorded and transcribed, and interview transcripts were accessible to the research team only. Detailed field notes were made after the calls and used to clarify comments made in the transcripts, but transcripts were not returned to participants for comment.
