## Appendix B for "The Role of Sepsis Care in Rural Emergency Departments: A Qualitative Study of Emergency Department User Perspectives"

**Supplemental Appendix B. Qualitative Interview Guide.**

### Qualitative Interview Guide

TELEViSED

TELEmedicine as a Virtual Intervention for Sepsis in Emergency Departments

**Topic:** How does provider-to-provider emergency department telehealth affect the care of rural sepsis patients?

*December 14, 2021*

**TABLE OF CONTENTS**

| | |
|---|---|
| *Project Summary* | *3* |
| *Interview Guide – Rural Physician or Advanced Practice Provider* | *4* |
| *Interview Guide – Rural Nurse* | *14* |
| *Interview Guide – Hub Physician* | *24* |

**TELEMEDICINE AS A VIRTUAL INTERVENTION FOR SEPSIS IN EMERGENCY DEPARTMENTS (TELEVISED)**

*Project Summary*

***Purpose:*** To elicit perspectives of rural physicians, advanced practice providers, nurses, and hub physician on the use of provider-to-provider telemedicine for the care of patients with sepsis in rural emergency departments.

***Population***: We will conduct interviews with the following individuals:

- Rural emergency physicians and advanced practice providers (n=15)
- Rural nurses (n=15)
- Hub physicians (n=5)

***Selection:*** Each site will be asked to identify the provider (MD vs PA/NP) and nurse who works the most clinical hours each month. From those contacts, we will select providers and nurses purposively to ensure representation of hospitals with different (1) telemedicine use (from quantitative data), (2) rurality (rural vs. frontier), (3) provider staffing patterns (physician vs. advanced practice provider), (4) local specialist availability, and (5) ED volume. These participants (providers and nurses) will be selected from the same 15 hospitals so that we can explore contextual differences in roles within hospitals. We will also interview 5 hub physicians, selected to ensure broad representation by (1) time in practice, (2) experience working in rural hospitals, and (3) duration working in the telemedicine hub. At the conclusion of interviews, any under-represented groups (e.g., females, frontier hospitals, family physicians covering the ED) will be intentionally recruited to supplement interviews prior to ending the study. In this way, our purposive sample will be focused on key informants most likely to give diverse opinions.

***Format:*** Interviews will be conducted by telephone over approximately 45 minutes each.

***Analysis:*** We will distribute anonymized transcripts to 2 reviewers. We will use inductive content analysis to identify themes related to each interview topic. Analysis will be based on a modified grounded theory analysis approach, in which we will allow themes to emerge from interview transcripts. We will engage in an open-focused coding process where each transcript will be systematically reviewed with investigators going sentence by sentence to identify codes. Codes that capture the main concepts will be identified and then compared across interviews using a process of “constant comparison” to identify overall themes around telemedicine use. Analysis will be conducted in parallel with ongoing interviews, thus allowing for newly emerging findings to be more fully explored, and interviews will continue until theme saturation is reached. Investigators will meet to review codes and themes until agreement is reached.

***Interview Guide – Rural Physician or Advanced Practice Provider***

****This interview guide has been annotated. Text that should be spoken is printed in **bold**. Questions are printed in **bold red**, while introduction is printed in **bold black**. Skip logic is included in <BRACKETS WITH CAPITAL LETTERS.> Optional probes are printed in un-bold red, and these probes should be used to elicit information that does not come out in the rest of your conversation.*

Introduction:

**Thank you for speaking with *me/us* today. As you know, we are conducting a research project to understand how people use emergency department telemedicine to care for patients with sepsis and how telemedicine impacts sepsis survival for those patients. As part of this project, we are talking with physicians and advanced practice providers working in rural emergency departments that use the eCARE service. Is it still all right that we talk today?**

**Is it all right with you that I record our interview today? We will keep your participation confidential, and we will only put de-identified information in any papers or reports that we write. Do you have any questions about that?**

**We also are compensating interview participants for your time talking with us today. At the end of the call, I will get your name and mailing address, so that we can mail you a check for $95 to your home address. Do you have any questions about that?**

**Just to confirm, you are still working at <HOSPITAL>, correct?**

**How long have you personally been in a role using the eCARE service?**

**This interview will take no more than 45 minutes. This is different from a survey—we're really hoping to hear about your experiences and stories of what you have experienced. We are looking for details to understand your feelings, perspectives, and observations, so feel free to share stories about your experiences. Do you have any questions before we get started?**

**For the purpose of the questions that I will be asking, have you personally used eCARE to take care of a sepsis patient in your hospital?**

> <If not, we can ask if there would be someone else at their site who they feel would be more suited to telling us about the use of eCARE for sepsis patients. Then we can terminate the interview.>

Question 1:

**I'd like to start by trying to understand how sepsis patients are treated in your emergency department. To do that, try to think back to a typical sepsis patient you treated recently. I'd like you to lead me through how a typical sepsis patient presents and how your team manages the care. I'm especially interested in your systems of care—screening processes, order sets, or any tools that you use to help your team care for sepsis patients. I don't want you to share any confidential information about the patient specifically, but I'm very interested in how that patient was treated in your emergency department.**

> We are really trying to understand how eCARE fits into the usual systems of care in participating EDs, so it's important that we understand what that system of care looks like. We want to probe to understand better WHO is performing different parts of that care, anyone locally who gets CALLED, the role of nurses in providing sepsis care, any decision support tools, order sets, screening systems, checklists, or anything else that they use in this hospital to help provide sepsis care better.

*<IF DESCRIPTION DOES NOT CLARIFY WHETHER E-CARE WAS USED, THEN ASK THE FOLLOWING QUESTION.>* **For this particular case, did you use eCARE to help you provide care for this patient?**

Question 2:

**Who is it that usually decides whether to consult eCARE for sepsis cases? Is it you or someone else on your team?**

> In this question, we're trying to understand whether the interviewee drives that decision or if someone else does. We are especially interested in whether there are protocols/expectations/guidance driving the use of eCARE. We also may start to hear in this question *reasons* for eCARE use—if that starts to come out, it is okay to finish that conversation here (otherwise, this information will be collected in Question 3).

Question 3:

*Modify this question as needed in response to who decides to consult eCARE:*

**What are some reasons why *you/your nurse* might choose to consult eCARE to help you take care of a sepsis case? Why?**

**When you've decided not to use eCARE for your sepsis patients, what are the reasons that you chose not to? Why not?**

> We would like to probe to make sure that we get a really comprehensive list to both of these prompts. We also want to try to elicit a detailed justification for how this decision whether to use eCARE is made. *If the second prompt does not elicit a rich response, consider using the following probing question:*

*Are there barriers to using eCARE, or things about the care of sepsis patients in particular that make eCARE less useful?*

In this question, we also want to understand whether eCARE helps with different phases of sepsis care. If those details do not come out in the answer, consider using the following prompts.

*To what extent does eCARE help you with any of the following parts of sepsis care:*

- *Sepsis identification or diagnosis;*
- *Sepsis treatment in your emergency department;*
- *Adherence with sepsis protocols you have in your hospital;*
- *Admission or transfer of your sepsis patients; or*
- *Anything else?*

Question 4:

**At what point in your sepsis patient encounter do you consider consulting eCARE, if you're going to? What drives that decision about timing?**

One of the facts we observe in the data is that eCARE is *very often* consulted AFTER the first hour of care in the ED. We are trying to understand why late consultation is so common, and if that has to do with the *reason* for consulting eCARE in the first place. Please probe to understand the issue of timing, who is activating eCARE, and how the reason for consultation influences timing of activating the network.

**Are there times when you consult eCARE earlier or later than that? Why?**

This question is ONLY a probe to try to understand to what extent timing of consultation is heterogeneous. If the practice for the individual is not uniform, we are most interested in **why not**.

Question 5:

**In what proportion of your sepsis patient encounters do you estimate that you use eCARE?**

This question is being asked for background so that you can use the response to understand the adequacy of the response to **Question 3**. If the interviewee gives you multiple reasons why telehealth is useful, then the consultation proportion is very low, then ask a probing follow-up question:

That surprises me. You told me about all the useful things that telehealth does to help you with sepsis care. I'm surprised you don't use it more.

If the interviewee gives you multiple barriers to telehealth use, but the consultation proportion is very high, then ask a probing follow-up question:

That surprises me. You told me about a number of barriers to telehealth being more useful for sepsis care. I'm surprised you use it so much.

These questions are both being used to make sure that our responses to **Question 3** are comprehensive.

Question 6:

**When you call eCARE for a sepsis patient, are there times when you disagree with what they are advising you to do? If so, how do you handle that?**

In this question, we are trying to understand ***to what extent telehealth threatens autonomy of the local care team***. We are also interested in how they view the participation of the telehealth provider as their ally. The interviewee may relate a story or example—follow-up that example with a question to understand to what extent local autonomy contributes to the decision to consult or the decision to adhere with eCARE recommendations.

We are also trying to understand local providers' adherence with recommendations. If there is an example of a disagreement over care, consider asking a follow-up question about how that was resolved:

It sounds like you didn't agree. Did you follow the recommendation from the eCARE provider or not?

Question 7:

**How do you think that having eCARE involved with sepsis care impacts the treatments that sepsis patients receive? How about their clinical outcomes, like survival or hospital length-of-stay?**

This question is asking the interviewee to "predict" how telehealth impacts:

- Specific elements of sepsis care; and
- Clinical outcomes.

Please use prompts to try to elicit answers to both of these.

**Are there other outcomes that you think eCARE might improve for sepsis patients?**

The following questions are really follow-up questions. They should only be used if these specific interventions did not come up in the primary answer.

*<Only ask this question if procedural support did not come up in the primary answer.>*

**To what extent does eCARE help you with the decision-making or completion of procedures for sepsis patients, such as intubation or central line placement?**

*<Only ask this question if medication/pharmacy support did not come up in the primary answer.>*

**To what extent does eCARE help you with medication selection or dosing in sepsis patients?**

Common answers to this prompt might include antibiotics selection or dosing or resuscitation medications (like vasopressors or intubation medications).

Question 8:

**Do you think there are particular sepsis patients who are more likely or less likely to benefit from eCARE consultation in your hospital? What are those groups and why?**

Question 9:

**The next questions focus on how others use the eCARE service.**

**What do other physicians or advanced practice providers in your group think about the use of eCARE for sepsis patients?**

**How about your nurses?**

In both of these questions, we're trying to elicit minority opinions. If, for instance, the interviewee tells us that eCARE service is amazing, would all of his/her partners and nurses agree? If not, why not?

**Do you think that your hospital's use of eCARE is typical for most eCARE hospitals, as it relates to sepsis care?**

Here, we are trying to understand how eCARE delivery might be different based on hospital characteristics. For instance, if the respondent is in a frontier hospital, perhaps there are characteristics of their health system that are unique to them that make them more or less likely to use the service. Similarly, there could be characteristics of larger hospitals that affect them similarly.

Question 10:

**To what extent do you learn about advances in sepsis care from eCARE encounters?**

The goal of this question is to understand how eCARE encounters contribute to ongoing medical training. Depending on the answers elicited, consider probing with some of the following questions:

If there is some learning, for instance, is eCARE ever consulted (during a patient encounter) for the purpose of learning?

Does this make it more or less likely that they will activate eCARE for the next encounter?

*<IF THE PARTICIPANT FEELS THAT THEY DO LEARN SOMETHING FROM THEIR ECARE ENCOUNTERS…>*

**How frequently do those lessons that you take from your encounters impact the care of future sepsis patients when you do not use eCARE? In what way—can you give me an example?**

We are interested in this question whether overall care is improving despite eCARE not being consulted on individual patients. Probing questions can ask how this knowledge influences future consults and how those consults standardize care across the members of the rural hospital staff.

Question 11:

**When we started this project, we expected that emergency department telemedicine would improve survival and length-of-stay for sepsis patients that came through the emergency department. That isn't really what we found—we found in almost 1300 patients that outcomes and care in those treated with eCARE were pretty similar to those who were not. In the next few questions, I'd like to share some of our findings to get your reactions.**

**First, does it surprise you that we didn't see better survival or shorter hospital stays in patients who had eCARE consulted?**

**What are some reasons why you think we might not have seen any differences?**

Question 12:

**We saw some pretty big differences in how often eCARE was used across hospitals and also among doctors and advanced practice providers within hospitals.  Some hospitals and providers used eCARE for the vast majority of their sepsis patients, and others used it rarely.  Does that surprise you?**

**What are some factors that you think might explain why some providers use eCARE for sepsis patients so much more than other people?**

The goal of this entire question is to elicit reasons for heterogeneity in how often eCARE is used.

Question 13:

**We've heard some providers tell us that sepsis care is much more standardized than care for some other diseases.  To what extent do you think that makes telehealth more or less important for sepsis care?**

Question 14:

**One of the other things we saw in our data was that eCARE is very frequently used in patients who need to be transferred for their sepsis care. Does that surprise you?  Why or why not?**

In this question, we want to understand the direction of causality: does eCARE *drive* transfer, or is that why eCARE is used.

*<If that does not come out in the answer to the prime* ***Question 14****, please ask the follow-up question.>*

**To what extent does the eCARE provider help you *decide* whether a patient needs to be transferred, and how often are you calling eCARE *because* you would like help getting a patient transferred?**

Question 15:

**Sepsis can be hard to diagnose and recognize.  How do you anticipate that sepsis recognition plays into the decision to use or not use eCARE?**

**Does eCARE ever help you recognize sepsis? Are there opportunities for telehealth to help you with the recognition and diagnosis of sepsis in the emergency department? How?**

> The goal of these questions is to understand how eCARE encounters contribute to improved sepsis recognition. If the respondent does not recognize that eCARE does help with diagnosis, then we want to probe to find ways that the service COULD help with recognition.

Question 16:

**You have experience using telehealth for a variety of problems in the ED. Are there things about the eCARE platform or service that you think would make it *more* useful for patients with sepsis in your emergency department? What would those things be and why?**

Question 17:

**That's all the questions that I have for you today. Do you have any other insights about telemedicine use for sepsis that I didn't ask about or that would help me understand how eCARE affects the care of sepsis patients in your hospital?**

**I am going to stop our recording now.**

**I mentioned at the beginning that we would be compensating you for your time during this interview. You will be receiving a check for $95 from the University of Iowa, mailed to your home address. For the purpose of receiving this compensation, can you please say and spell your name?**

**What is your mailing address?**

**Thank you very much for your time today. We really value your insight and perspective. If you have any other thoughts that come up after the interview, please feel free to reach out again. When this project is finished, we will be writing a paper about telemedicine use for sepsis, and we will share that with the eCARE hub. You should be receiving a check to your home address within the next 6 weeks. Do you have any other questions for me?**

***Interview Guide – Rural Nurse***

****This interview guide has been annotated. Text that should be spoken is printed in **bold**. Questions are printed in **bold red**, while introduction is printed in **bold black**. Skip logic is included in <BRACKETS WITH CAPITAL LETTERS.> Optional probes are printed in un-bold red, and these probes should be used to elicit information that does not come out in the rest of your conversation.*

Introduction:

**Thank you for speaking with *me/us* today. As you know, we are conducting a research project to understand how people use emergency department telemedicine to care for patients with sepsis and how telemedicine impacts sepsis survival for those patients. As part of this project, we are talking with nurses working in rural emergency departments that use the eCARE service. Is it still all right that we talk today?**

**Is it all right with you that I record our interview today? We will keep your participation confidential, and we will only put de-identified information in any papers or reports that we write. Do you have any questions about that?**

**We also are compensating interview participants for your time talking with us today. At the end of the call, I will get your name and mailing address, so that we can mail you a check for $95 to your home address. Do you have any questions about that?**

**Just to confirm, you are still working at <HOSPITAL>, correct?**

**How long have you personally been in a role using the eCARE service?**

**This interview will take no more than 45 minutes. This is different from a survey—we're really hoping to hear about your experiences and stories of what you have experienced. We are looking for details to understand your feelings, perspectives, and observations, so feel free to share stories about your experiences. Do you have any questions before we get started?**

**For the purpose of the questions that I will be asking, have you personally used eCARE to take care of a sepsis patient in your hospital?**

<If not, we can ask if there would be someone else at their site who they feel would be more suited to telling us about the use of eCARE for sepsis patients. Then we can terminate the interview.>

Question 1:

**I'd like to start by trying to understand how sepsis patients are treated in your emergency department. To do that, try to think back to a recent sepsis patient you treated. I'd like you to lead me through how that case presented and how your team managed the care. I'm especially interested in your systems of care—screening processes, order sets, or any tools that you use to help your team care for sepsis patients. I don't want you to share any confidential information about the patient specifically, but I'm very interested in how that patient was treated in your emergency department.**

> We are really trying to understand how eCARE fits into the usual systems of care in participating EDs, so it's important that we understand what that system of care looks like. We want to probe to understand better WHO is performing different parts of that care, anyone locally who gets CALLED, the role of nurses in providing sepsis care, any decision support tools, order sets, screening systems, checklists, or anything else that they use in this hospital to help provide sepsis care better.

*<IF DESCRIPTION DOES NOT CLARIFY WHETHER E-CARE WAS USED, THEN ASK THE FOLLOWING QUESTION.>* **For this particular case, did you use eCARE to help you provide care for this patient?**

Question 2:

**Who is it that usually decides whether to consult eCARE? Is it you or someone else on your team?**

> In this question, we're trying to understand whether the interviewee drives that decision or if someone else does. We are especially interested in whether there are protocols/expectations/guidance driving the use of eCARE. We also may start to hear in this question *reasons* for eCARE use—if that starts to come out, it is okay to finish that conversation here (otherwise, this information will be collected in Question 3).

Question 3:

*Modify this question as needed in response to who decides to consult eCARE:*

**What are some reasons why *you/a provider* might choose to consult eCARE to help you take care of a *particular* sepsis case? Why?**

> We are really looking for nursing-specific reasons. If the answer does not comment on nursing reasons, then probe with: Are there specific nursing reasons why you might consult eCARE?
>
> We have heard previously that documentation may be one reason that eCARE is used. It is okay to use this as a prompt/example, and we are also interested in other uses of eCARE or reasons for activating the service.

**When the treatment team has decided not to use eCARE for your sepsis patients, what are the reasons that you chose not to? Why not?**

We would like to probe to make sure that we get a really comprehensive list to both of these prompts. We also want to try to elicit a detailed justification for how this decision whether to use eCARE is made. *If the second prompt does not elicit a rich response, consider using the following probing question:*

*Are there barriers to using eCARE, or things about the care of sepsis patients in particular that make eCARE less useful?*

In this question, we also want to understand whether eCARE helps with different phases of sepsis care. If those details do not come out in the answer, consider using the following prompts.

*To what extent does eCARE help you with any of the following parts of sepsis care:*

- *Sepsis identification or diagnosis;*
- *Sepsis treatment in your emergency department;*
- *Adherence with sepsis protocols you have in your hospital;*
- *Admission or transfer of your sepsis patients*
- *Documentation; or*
- *Anything else?*

Question 4:

**At what point in your patient encounter do you or others on the treatment team consider consulting eCARE, if you're going to? What drives that decision about timing?**

One of the facts we observe in the data is that eCARE is *very often* consulted AFTER the first hour of care in the ED. We are trying to understand why late consultation is so common, and if that has to do with the *reason* for consulting eCARE in the first place. Please probe to understand the issue of timing, who is activating eCARE, and how the reason for consultation influences timing of activating the network.

**Are there times when you decide to consult eCARE earlier or later than that? Why?**

This question is ONLY a probe to try to understand to what extent timing of consultation is heterogeneous. If the practice for the individual is not uniform, we are most interested in **why not**.

Question 5:

**In what proportion of your sepsis patient encounters do you estimate that sepsis patients you care for in your emergency department have eCARE consulted as part of their care?**

This question is being asked for background so that you can use the response to understand the adequacy of the response to **Question 3**. If the interviewee gives you multiple reasons why telehealth is useful, then the consultation proportion is very low, then ask a probing follow-up question:

That surprises me. You told me about all the useful things that telehealth does to help you with sepsis care. I'm surprised you don't use it more.

If the interviewee gives you multiple barriers to telehealth use, but the consultation proportion is very high, then ask a probing follow-up question:

That surprises me. You told me about a number of barriers to telehealth being more useful for sepsis care. I'm surprised you use it so much.

These questions are both being used to make sure that our responses to **Question 3** are comprehensive.

Question 6:

**When you call eCARE for a sepsis patient, are there times when there is disagreement about whether to follow their recommendations? How does your team handle that?**

In this question, we are trying to understand ***to what extent telehealth threatens autonomy of the local care team***. We are also interested in how they view the participation of the telehealth provider as their ally. The interviewee may relate a story or example—follow-up that example with a question to understand to what extent local autonomy contributes to the decision to consult or the decision to adhere with eCARE recommendations.

We are also trying to understand local providers' adherence with recommendations. If there is an example of a disagreement over care, consider asking a follow-up question about how that was resolved:

It sounds like your team didn't agree. Did your team follow the recommendation from the eCARE provider?

Question 7:

**How do you think that having eCARE involved with sepsis care impacts the treatments that sepsis patients receive? How about their clinical outcomes, like survival or hospital length-of-stay?**

This question is asking the interviewee to "predict" how telehealth impacts:

- Specific elements of sepsis care; and
- Clinical outcomes.

Please use prompts to try to elicit answers to both of these.

**Are there other outcomes that you think eCARE might improve for sepsis patients?**

The following questions are really follow-up questions. They should only be used if these specific interventions did not come up in the primary answer.

*<Only ask this question if procedural support did not come up in the primary answer.>*

**To what extent does eCARE help your team with the *decision-making* or *completion* of <u>procedures</u> for sepsis patients, such as intubation or central line placement?**

*<Only ask this question if medication/pharmacy support did not come up in the primary answer.>*

**To what extent does eCARE help you with medication selection or dosing in sepsis patients?**

Common answers to this prompt might include antibiotics selection or dosing or resuscitation medications (like vasopressors or intubation medications).

<u>Question 8:</u>

**Do you think there are certain types of sepsis patients who are more likely or less likely to benefit from eCARE consultation in your hospital? What are those groups and why?**

<u>Question 9:</u>

**The next questions focus on how <u>others</u> use the eCARE service.**

**What do physicians or advanced providers at your hospital think about the use of eCARE for sepsis patients?**

**Do you see differences in eCARE use between physicians and advanced practice providers in your hospital? If so, why do you think that is?**

**What do your nursing colleagues think about the use of eCARE?**

In each of these questions, we're trying to elicit minority opinions. If, for instance, the interviewee tells us that eCARE service is amazing, would all of his/her partners and nurses agree? If not, why not?

Question 10:

**To what extent do you learn about advances in sepsis care from eCARE encounters?**

The goal of this question is to understand how eCARE encounters contribute to ongoing medical training. Depending on the answers elicited, consider probing with some of the following questions:

If there is some learning, for instance, is eCARE ever consulted for the purpose of learning?

Does this make it more or less likely that they will activate eCARE for the next encounter?

*<IF THE PARTICIPANT FEELS THAT THEY DO LEARN SOMETHING FROM THEIR ECARE ENCOUNTERS…>*

**How frequently do those lessons that you take from your encounters impact the care of future sepsis patients when you do not use eCARE? In what way—can you give me an example?**

We are interested in this question whether overall care is improving despite eCARE not being consulted on individual patients. Probing questions can ask how this knowledge influences future consults and how those consults standardize care across the members of the rural hospital staff.

Question 11:

**When we started this project, we expected that emergency department telemedicine would improve survival and length-of-stay for sepsis patients that came through the emergency department. That isn't really what we found—we found in almost 1300 patients that outcomes and care were pretty similar. In the next few questions, I'd like to share some of our findings to get your reactions.**

**First, does it surprise you that we didn't see better survival or shorter hospital stays in patients who had eCARE consulted?**

**What are some reasons why you think we might not have seen any differences?**

Question 12:

**We saw some pretty big differences in how often eCARE was used across hospitals and also among doctors and advanced practice providers within hospitals. Some rural hospitals used eCARE for the vast majority of their sepsis patients, and others used it rarely. Does that surprise you?**

**What are some factors that you think might explain why some people use eCARE for sepsis patients so much more than other people?**

The goal of this entire question is to elicit reasons for heterogeneity in how often eCARE is used.

Question 13:

**We've heard some people tell us that sepsis care is much more standardized than care for some other diseases. To what extent do you think that makes telehealth more or less important for sepsis care?**

Question 14:

**One of the other things we saw in our data was that eCARE is very frequently used in patients who need to be transferred for their sepsis care. Does that surprise you? Why or why not?**

In this question, we want to understand the direction of causality: does eCARE *drive* transfer, or is that why eCARE is used.

*<If that does not come out in the answer to the prime* ***Question 14****, please ask the follow-up question.>*

**To what extent does the eCARE provider help you *decide* whether a patient needs to be transferred, and how often are you calling eCARE *because* you would like help getting a patient transferred?**

Question 15:

**Sepsis can be hard to diagnose and recognize. How do you anticipate that sepsis recognition plays into the decision to use or not use eCARE?**

**Does eCARE every help you recognize sepsis? Are there opportunities for telehealth to help you with the recognition and diagnosis of sepsis in the emergency department? How?**

> The goal of these questions is to understand how eCARE encounters contribute to improved sepsis recognition. If the respondent does not recognize that eCARE does help with diagnosis, then we want to probe to find ways that the service COULD help with recognition.

Question 16:

**You have experience using telehealth for a variety of problems in the ED. Are there things about the eCARE platform or service that you think would make it *more* useful for patients with sepsis in your emergency department? Would would those things be and why?**

Question 17:

**That's all the questions that I have for you today. Do you have any other insights about telemedicine use for sepsis that I didn't ask about or that would help me understand how eCARE affects the care of sepsis patients in your hospital?**

**I am going to stop our recording now.**

**I mentioned at the beginning that we would be compensating you for your time during this interview. You will be receiving a check for $95 from the University of Iowa, mailed to your home address. For the purpose of receiving this compensation, can you please say and spell your name?**

**What is your mailing address?**

**Thank you very much for your time today. We really value your insight and perspective. If you have any other thoughts that come up after the interview, please feel free to reach out again. When this project is finished, we will be writing a paper about telemedicine use for sepsis, and we will share that paper with the eCARE hub. Do you have any other questions for me?**

This page intentionally left blank.

***Interview Guide – Hub Physician***

****This interview guide has been annotated. Text that should be spoken is printed in **bold**. Questions are printed in **bold red**, while introduction is printed in **bold black**. Skip logic is included in <BRACKETS WITH CAPITAL LETTERS.> Optional probes are printed in un-bold red, and these probes should be used to elicit information that does not come out in the rest of your conversation.*

Introduction:

**Thank you for speaking with *me/us* today. As you know, we are conducting a research project to understand how people use emergency department telemedicine to care for patients with sepsis and how telemedicine impacts sepsis survival for those patients. As part of this project, we are talking with physicians working in the eCARE hub. Is it still all right that we talk today?**

**Is it all right with you that I record our interview today? We will keep your participation confidential, and we will only put de-identified information in any papers or reports that we write. Do you have any questions about that?**

**We also are compensating interview participants for your time talking with us today. At the end of the call, I will get your name and mailing address, so that we can mail you a check for $95 to your home address. Do you have any questions about that?**

**Just to confirm, you are still working in the eCARE hub, correct?**

**How long have you been working at eCARE?**

**This interview will take no more than 45 minutes. This is different from a survey—we're really hoping to hear about your experiences and stories of what you have experienced. We are looking for details to understand your feelings, perspectives, and observations, so feel free to share stories about your experiences. Do you have any questions before we get started?**

**For the purpose of the questions that I will be asking, have you personally participated in eCARE calls to help take care of a sepsis patient?**

> <If not, we can ask if there would be someone else at the hub who they feel would be more suited to telling us about the use of eCARE for sepsis patients. Then we can terminate the interview.>

Question 1:

**I'd like to start by trying to understand how sepsis patients are treated with the participation of eCARE. To do that, try to think back to a recent sepsis patient you treated through your video service. I'd like you to lead me through how that case presented and how your team helped to provide care. I'm especially interested in how eCARE integrates into the local system of care—screening processes, order sets, or any tools that you use to help your team care for sepsis patients. I don't want you to**

**share any confidential information about the patient specifically, but I'm very interested in how that patient was treated.**

We are really trying to understand how eCARE fits into the usual systems of care in participating EDs, so it's important that we understand what that system of care looks like. We want to probe to understand better WHO is performing different parts of that care, anyone locally who gets CALLED, the role of nurses in providing sepsis care, any decision support tools, order sets, screening systems, checklists, or anything else that they use in this hospital to help provide sepsis care better.

Question 2:

**Can you tell from your interactions who consulted eCARE? If so, how can you tell? Who is it that usually decides whether to consult eCARE?**

We are especially interested in whether there are protocols/expectations/guidance driving the use of eCARE. We also may start to hear in this question *reasons* for eCARE use—if that starts to come out, it is okay to finish that conversation here (otherwise, this information will be collected in Question 3).

Question 3:

**What are some main reasons why one of your customers might choose to consult eCARE to help you take care of a *particular* sepsis case? Why?**

**Are you aware of cases when your customers choose not to use eCARE? What are some of the reasons it might not be used?**

In this question, we also want to understand whether eCARE helps with different phases of sepsis care. If those details do not come out in the answer, consider using the following prompts.

*To what extent does eCARE help you with any of the following parts of sepsis care:*

- *Sepsis identification or diagnosis;*
- *Sepsis treatment in your emergency department;*
- *Adherence with sepsis protocols you have in your hospital;*
- *Admission or transfer of your sepsis patients*
- *Documentation; or*
- *Anything else?*

Question 4:

**At what point in the rural patient encounter is eCARE usually consulted? What do you think drives that decision about timing?**

One of the facts we observe in the data is that eCARE is *very often* consulted AFTER the first hour of care in the ED. We are trying to understand why late consultation is so common, and if that has to do with the *reason* for consulting eCARE in the first place. Please probe to understand the issue of timing, who is activating eCARE, and how the reason for consultation influences timing of activating the network.

**How does timing of consultation affect the manner in which you are able to contribute to the care of a patient?**

Here we are trying to understand how recommendations or involvement might be different in early versus late consultation.

Question 5:

**Do you have the sense there is disagreement between the hub staff and the rural hospital treatment team about how to treat an individual sepsis patient? How do you handle that?**

We are trying to understand local providers' adherence with recommendations. If there is an example of a disagreement over care, consider asking a follow-up question about how that was resolved:

What was the underlying reason you disagreed?

**Are there ever times you make a recommendation that is not followed? If so, tell me about that.**

Question 6:

**How do you think that having eCARE involved with sepsis care impacts the treatments that sepsis patients receive? How about their clinical outcomes, like survival or hospital length-of-stay?**

This question is asking the interviewee to "predict" how telehealth impacts:

- Specific elements of sepsis care; and
- Clinical outcomes.

Please use prompts to try to elicit answers to both of these.

**Are there other outcomes that you think eCARE might improve for sepsis patients?**

The following questions are really follow-up questions. They should only be used if these specific interventions did not come up in the primary answer.

*<Only ask this question if procedural support did not come up in the primary answer.>*

**To what extent does eCARE help with procedures for sepsis patients, such as intubation or central line placement?**

*<Only ask this question if medication/pharmacy support did not come up in the primary answer.>*

**To what extent does eCARE help you with medication selection or dosing in sepsis patients?**

Common answers to this prompt might include antibiotics selection or dosing or resuscitation medications (like vasopressors or intubation medications).

Question 7:

**Do you think there are particular sepsis patients who are more likely or less likely to benefit from eCARE consultation? What are those groups and why?**

Question 8:

**Has there ever been a time when you have been able to help recognize or diagnose sepsis? If so, tell me about a time when you were able to help a rural provider recognize or diagnose sepsis.**

*<IF PARTICIPANT ANSWERS YES>*

**How or why do you think you were able to recognize that condition differently than the local provider?**

Question 9:

**When we started this project, we expected that emergency department telemedicine would improve survival and length-of-stay for sepsis patients that came through the emergency department. That isn't really what we found—we found in almost 1300 patients that outcomes and care were pretty similar. In the next few questions, I'd like to share some of our findings to get your reactions.**

**First, does it surprise you that we didn't see better survival or shorter hospital stays in patients who had eCARE consulted?**

**What are some reasons why you think we might not have seen any differences?**

Question 10:

**We saw some pretty big differences in how often eCARE was used across hospitals and also among doctors and advanced practice providers within hospitals. Some hospitals and providers used eCARE for the vast majority of their sepsis patients, and others used it rarely. Does that surprise you?**

**What are some factors that you think might explain why some providers or nurses in rural hospitals use eCARE for sepsis patients so much more than other people?**

The goal of this entire question is to elicit reasons for heterogeneity in how often eCARE is used.

Question 11:

**We've heard some providers tell us that sepsis care is much more standardized than care for some other diseases. To what extent do you think that makes telehealth more or less important for sepsis care?**

Question 12:

**One of the other things we saw in our data was that eCARE is very frequently used in patients who need to be transferred for their sepsis care. Does that surprise you? Why or why not?**

In this question, we want to understand the direction of causality: does eCARE *drive* transfer, or is that why eCARE is used.

*<If that does not come out in the answer to the prime **Question 12**, please ask the follow-up question.>*

**To what extent does eCARE help a local provider *decide* whether a patient needs to be transferred, and how is eCARE consulted *because* a local provider would like help getting a patient transferred?**

Question 13:

**You have a lot of experience providing telehealth for a variety of problems in rural EDs. Are there things about the eCARE platform or service that you think would make it *more* useful for patients with sepsis in rural emergency departments? What would those things be and why?**

Question 14:

**That's all the questions that I have for you today. Do you have any other insights about telemedicine use for sepsis that I didn't ask about or that would help me understand how eCARE affects the care of sepsis patients?**

**I am going to stop our recording now.**

**I mentioned at the beginning that we would be compensating you for your time during this interview. You will be receiving a check for $95 from the University of Iowa, mailed to your home address. For the purpose of receiving this compensation, can you please say and spell your name?**

**What is your mailing address?**

**Thank you very much for your time today. We really value your insight and perspective. If you have any other thoughts that come up after the interview, please feel free to reach out again. When this project is finished, we will be writing a paper about telemedicine use for sepsis, and we will share that with the eCARE hub. Do you have any other questions for me?**
